# Acceptable Performance of the Abbott ID NOW Among Symptomatic Individuals with Confirmed COVID-19

**DOI:** 10.1101/2020.12.24.20248786

**Authors:** William Stokes, Byron M. Berenger, Takshveer Singh, Ifueko Adeghe, Angela Schneider, Danielle Portnoy, Teagan King, Brittney Scott, Kanti Pabbaraju, Sandy Shokoples, Anita A. Wong, Kara Gill, LeeAnn Turnbull, Jia Hu, Graham Tipples

**Affiliations:** Public Health Laboratory, Alberta Precision Laboratories, Alberta, Canada; Department of Pathology and Laboratory Medicine, University of Alberta, Alberta, Canada; Division of Infectious Diseases, Department of Medicine, University of Alberta, Edmonton, Alberta; Department of Pathology and Laboratory Medicine, University of Calgary, Calgary, Alberta, Canada; Cumming School of Medicine, University of Calgary, Calgary, Alberta; Faculty of Medicine & Dentistry, University of Alberta, Edmonton, Alberta; Department of Community Health Sciences, University of Calgary, Calgary, Alberta, Canada; Public Health, Alberta Health Services, Alberta, Canada; Li Ka Shing Institute of Virology, University of Alberta, Edmonton, Alberta, Canada

**Keywords:** ID NOW, rapid SARS-CoV-2 test, COVID-19 diagnostics

## Abstract

Point of care diagnostic tests for SARS-CoV-2, such as the ID NOW, have great potential to help combat the COVID-19 pandemic. The ID NOW is approved by the United States Food and Drug Administration (FDA) for the detection of SARS-CoV-2 in symptomatic individuals within the first 7 days of symptom onset for COVID-19 if tested within 1 hour of specimen collection. However, clinical data on the performance of the ID NOW is limited, with many studies deviating from the manufacturer’s instructions and/or having small sample size.

**METHODS:** Adults with COVID-19 in the community or hospital were recruited into the study. Paired throat swabs were collected, with one throat swab transported immediately in an empty sterile tube to the laboratory for ID NOW testing, and the other transported in universal transport media and tested by an in-house SARS-CoV-2 RT-PCR assay targeting the E-gene. Positive percent agreement (PPA) was calculated.

**RESULTS:** 133 individuals were included in the study. 129 samples were positive on either the ID NOW and/or RT-PCR. Assuming any positive result on either assay represents a true positive, PPA of the ID NOW compared to RT-PCR with 95% confidence intervals was 89.1% [82.0% - 94.1%] and 91.6% [85.1% - 95.9%], respectively. When analyzing individuals with symptoms ≤ 7 days and who had the ID NOW performed within an hour, ID NOW PPA increased to 98.2%.

**DISCUSSION:** In this study, SARS-CoV-2 results from the ID NOW were reliable, especially when testing was adhered to manufacturer’s recommendations.

## INTRODUCTION

The ID NOW (Abbott, Illinois, United States) is approved by the United States Food and Drug Administration (FDA) for the detection of severe acute respiratory syndrome-coronavirus-2 (SARS-CoV-2) in individuals who are within the first 7 days of symptom onset. The assay targets a region of the RNA-dependent RNA polymerase (RdRp) gene of SARS-CoV-2 for using isothermal amplification and subsequent detection of fluorescence, with results available in under 15 minutes. Clinical specimens approved for testing include nasal, throat, and nasopharyngeal swabs. These specimens must be tested on the Abbott ID NOW either immediately or within 1 hour of collection if stored in a clean, unused tube. Specimens placed in viral/universal transport media (UTM) are not valid for testing on the Abbott ID NOW.^1^

Current limitations of the Abbott ID NOW include the lack of strong data to determine its effectiveness in detecting SARS-CoV-2 in clinical settings. To date, the studies used to obtain FDA approval have not used clinical specimens. These studies have demonstrated that the limit of detection of the Abbott ID NOW is similar to other nucleic acid amplification tests at approximately 125 genome equivalents/mL. Of the clinical studies reported in the literature, the Abbott ID NOW has been shown to have excellent specificity (∼100%) but sensitivity varies widely between studies (48.0% - 94.1%).^2-17^ In addition, many of these studies suffer from poor study design such as comparing nasopharyngeal to nasal specimens, deviating from the product insert or having major delays in testing specimens on the ID NOW. Only two studies adhered to the FDA’s recommendations for ID NOW testing, and both studies had a small sample size (<20 samples positive for SARS-CoV-2).^16,17^

We sought to assess the positive percent agreement (PPA) of the ID NOW by comparing its performance to an in-house validated real-time reverse transcriptase (RT-PCR) among individuals with recently confirmed COVID-19 while adhering as closely as possible to the manufacturers recommendations. We also tested the accuracy of the ID NOW with samples taken from asymptomatic individuals at low risk for COVID-19 (i.e. no exposures) and on retrospective clinical samples previously positive for common respiratory pathogens.

## METHODS

### Testing individuals with confirmed COVID-19

Community and hospitalized individuals within the Calgary and Edmonton Health Zones of Alberta, Canada, who recently tested positive for SARS-CoV-2 at Alberta Precision Laboratories (APL) and confirmed as cases by Alberta Health Services (AHS) Public Health were recruited. Diagnostic testing was performed by a Health Canada approved SARS-CoV-2 assay or a lab developed real time RT-PCR assay (see below for details). Participants were identified by an AHS Public Health confirmed case list. Oral consent by phone was obtained for collection of samples in the participant’s home or in a hospital (if hospitalized). Individuals under the age of 18 and individuals in supportive or congregate living facilities were excluded. Eligible patients who consented to the study were recruited to have two throat swabs collected by trained healthcare professionals within their homes or inpatient unit. The University of Calgary Research Ethics board approved this study (REB20-444).

Individuals were asked to confirm their symptoms and date of symptom onset at the time of study swab collection. Healthcare workers performing the collection were given instructions on how to perform throat swabs using the ClassiqSwabs (COPAN Diagnostics, California, United States) and the throat swab provided in the ID NOW testing kits (Abbott, Illinois, United States).^18^ Throat swabs were collected from both sides of the oropharynx and the posterior pharyngeal wall under the uvula. Throat swabs were collected approximately one minute apart and the order in which throat swabs were collected was recorded.

For each paired throat swab, one was placed into a dry 15mL conical centrifuge tube (Fisher Scientific, MA, United States) for ID NOW testing and the other into a tube containing UTM (COPAN) for RT-PCR testing. Samples were transported to the APL Public Health Laboratory as quickly as possible, at room temperature, and tested upon receipt. Testing on the ID NOW instrument was done immediately upon receipt as per manufacturer’s instructions. Throat swabs in UTM collected for RT-PCR testing were stored at 4°C and tested within 72 hours. Two hundred microliters of UTM were extracted on the MagMAX Express-96 or Kingfisher Flex (ABI) using the MagMAX-96 Viral RNA Isolation Kit (ThermoFisher) or the PurePrep Pathogen Kit (MolGen) according to manufacturer’s instructions, and eluted into a volume of 110ul. RT-PCR testing included an assay targeting the envelope (E)-gene of SARS-CoV-2 developed and validated at APL, and the Cobas® SARS-CoV-2 (Roche Diagnostics, Indianapolis, IN) test on the Cobas 6800 instrument.^19^

For our lab-developed test, the samples were considered positive for SARS-CoV-2 when the cycle threshold (Ct) value was <35. If the Ct was ≥35, amplification from the same eluate was repeated in duplicate and was considered positive if at least 2/3 results had a Ct <41. Testing for SARS-CoV-2 on the Cobas 6800 instrument was performed according to manufacturer’s instructions. For the Cobas SARS-CoV-2 test, a positive result was defined as 2/2 targets positive or 1 or more targets positive in duplicate. If 1/2 targets were positive and duplicate testing was negative, the result was considered indeterminate.

For discrepant results, the specimens were retested in triplicate with our lab developed RT-PCR test and in triplicate with the N2 assay from the CDC 2019-Novel Coronavirus (2019-nCoV) Real-Time RT-PCR Diagnostic Panel using the UltraPlex 1-Step Toughmix (Quantabio, MA, USA).^20^ If still negative, the test was run on the Cobas 6800. PPA was calculated with Clopper-Pearson 95% confidence intervals. Statistical analysis was performed using Pearson Chi-squared for categorical variables and t-test for continuous variables using STATA (version 14.1).

### Negative samples and retrospective samples containing other respiratory viruses

Two throat swabs were collected from asymptomatic individuals at low risk of having COVID-19 (no recent travel, no exposures). One throat swab was tested immediately (<2 minutes) on the ID NOW instrument. The other throat swab was tested by RT-PCR as explained above. To assess for cross-reactivity, retrospective samples containing high concentrations of various respiratory viruses, stored in UTM, were tested by aliquoting 400uL of sample into the ID NOW blue specimen receiver. These samples were previously tested by the NxTAG® Respiratory Pathogen Panel (Luminex, Tx, United States) or the CDC influenza SARS-CoV-2 multiplex assay. The ability of the ID NOW to process this volume of UTM was confirmed by testing four retrospective positive SARS-CoV-2 samples, in UTM, and showing that they could be detected (data not shown, Ct values ranging from 21 - 28).

## RESULTS

One hundred and fifty-two patients were recruited for this study. Fourteen individuals were asymptomatic at the time of COVID-19 diagnosis and at time of study collection and were therefore excluded from analysis (sub-analysis provided in the supplementary material). The date of symptom onset for two individuals was not captured at time of consent and they could not be reached later for clarification, and therefore excluded. Three samples were excluded as samples were lost for RT-PCR testing (all were ID NOW positive). Symptom details were not recorded for two individuals but they were still included in the analysis as symptom onset was known and they were still symptomatic at the time of collection. This resulted in the inclusion of 133 individuals in our analysis and their characteristics are provided in Table 1.

**Table 1:** Patient characteristics (N=133).

| Characteristic |  |
| --- | --- |
| Male gender | 36.8 |
| Mean age in years (median, range) | 42.9 (40.4, 19.0 – 92.7) |
| Inpatient/Hospitalized at time of collection | 9.0% |
| ID NOW throat swab collected first | 78.0% |
| Mean time from sample collection to ID NOW testing (median, range), N=133 | 54.8 min (54, 20 – 233) |
| Samples tested on ID NOW within 1 hour from collection | 62.4% |
| Mean time from starting ID NOW test to confirming positive result (median, range), N=106 | 2.4 min (2, 1 – 8 min) |
| Mean duration of symptoms from collection date (median, range) | 6.9 days (7, 1 – 17 days) |
| Individuals with symptom duration $\leq 7$ days from collection date | 75.2% |
| Individuals with symptoms $\leq 7$ days and ID NOW test conducted within an hour from collection, (N=62) | 45.1% |

All individuals had symptoms at the time of collection and the majority (91.0%) were from the community. For the 131 individuals with recorded symptoms, cough was the most frequent symptom (40.5%), followed by pharyngitis (31.3%), fevers/chills (30.5%), headache (24.4%), nasal congestion (23.7%), anosmia (22.9%), malaise (21.4%), myalgias (21.4%), ageusia (19.1%), shortness of breath (13.0%), rhinorrhea (11.5%), nausea/vomiting (3.8%) and other including chest pain, diarrhea, loss of appetite or arthralgias (7.6%).

Mean duration of symptoms at the time of collection was 6.9 (median=7, range=1 – 17) days. Seventy-five percent (n=100) of individuals were within the 7-day symptom onset window, 62.4% (n=83) of individuals had their samples tested on the ID NOW within 1 hour from collection, and 46.6% (N=62) met the regulatory agency approved criteria for testing on the ID NOW (symptom onset ≤ 7 days and ID NOW test conducted within an hour from collection). The mean E-gene Ct value for positive results from the RT-PCR assay was 30.9 (median=31.0, range=16.4 – 37.9).

Of the 133 samples, 96 were positive on both the ID NOW and RT-PCR (Table 2). Assuming any positive result represents a true positive, the PPA of the ID NOW compared to RT-PCR with 95% confidence intervals was found to be 89.1% [82.0% - 94.1%] and 91.6% [85.1% - 95.9%], respectively (Table 3). There were 13 false negatives on the ID NOW instrument, with 11/13 (91.7%) having Ct value > 30 on RT-PCR (or indeterminate on the Cobas 6800) and 6/13 (41.7%) with Ct values > 37 on RT-PCR (or indeterminate on the Cobas 6800) (Table 4). Eight of the 13 false negative samples (61.5%) were from individuals within 7 days symptoms onset, four (30.7%) were samples that were tested on the ID NOW within an hour from collection, and only one (7.7%) was from an individual with symptoms ≤ 7 days and had the ID NOW performed within an hour. Samples tested on the ID NOW instrument were more likely to be positive if the sample was tested within one hour from collection (p=0.031) and from individuals who had lower Ct values on RT-PCR testing (p<0.001) (Table 5). RT-PCR samples were more likely to be positive if the sample was tested on individuals with symptoms ≤ 7 days (Table S2).

**Table 2:** Results of ID NOW and RT-PCR in symptomatic COVID-19 patients (N=133).

|  |  | RT-PCR |  |
| --- | --- | --- | --- |
|  |  | Positive | Negative |
| ID NOW | Positive | 96 | 10 |
|  | Negative | 13 | 14 |

**Table 3:** Positive percent agreement (PPA) between ID NOW and RT-PCR in symptomatic COVID-19 patients (N=133). PPA calculated assuming any positive is a true positive.

|  | PPA [95% CI] |
| --- | --- |
| ID NOW | 89.1% [82.0% – 94.1%] |
| RT-PCR | 91.6% [85.1% - 95.9%] |

**Table 4:** Details on the ID NOW negative, RT-PCR positive results (N=14).

| Sample Number | Ct value from RT-PCR | Days from symptom onset at time of collection | Symptoms | Time from sample collection to ID NOW testing (min) |
| --- | --- | --- | --- | --- |
| 1 | 33.2 | 7 | Pharyngitis, cough | 28 |
| 2 | 37.2 | 5 | Cough, malaise, myalgias, fever | 63 |
| 3 | 35.0 | 5 | Pharyngitis, anosmia | 79 |
| 4 | 34.3 | 6 | Pharyngitis, cough, malaise, fever | 63 |
| 5 | Indeterminate | 6 | Pharyngitis, malaise, cough, fever, myalgia, anosmia, ageusia | 64 |
| 6 | 31.7 | 6 | Rhinorrhea, fever | 94 |
| 7 | 33.8 | 7 | Pharyngitis, cough | 66 |
| 8 | 37.0 | 7 | Headache, decreased appetite, anosmia, ageusia | 72 |
| 9 | 36.8 | 8 | Malaise | 37 |
| 10 | 29.8 | 9 | Sore throat | 68 |
| 11 | 37.8 | 10 | Pharyngitis, myalgias | 78 |
| 12 | 37.8 | 11 | Shortness of breath | 36 |
| 13 | 37.8 | 12 | Cough, anosmia | 39 |
| MEAN (median) | 35.2 (35.9) | 7.6 (7) | N/A | 60.5 (64) |

Only one false negative sample (coloured in grey) was from an individual who had duration of symptoms $\leq 7$ days at time of collection and had their sample tested within 1 hour from collection.

**Table 5:** Characteristics between ID NOW negative and ID NOW positive samples (N=152).

|  | **ID NOW**<br>**negative**<br>**(N=27)** | **ID NOW positive**<br>**(N=106)** | **P-value** |
| --- | --- | --- | --- |
| **Sample tested within one hour from collection** | 44.4% | 67.0% | 0.031 |
| **Mean duration of symptoms** | 7.4 days | 6.8 days | 0.280 |
| **Symptoms ≤ 7 days at collection** | 63.0% | 78.3% | 0.099 |
| **Mean age** | 41.4 | 43.3 | 0.590 |
| **Mean Ct value** | 35.2 | 30.3 | <0.001 |
| **Throat swab tested on ID NOW collected first** | 77.8% | 78.1% | 0.972 |
| **Hospitalized** | 11.1% | 8.5% | 0.671 |
| **Male gender** | 44.4% | 34.9 | 0.359 |

When tested in triplicate using RT-PCR followed by triplicate testing by the CDC method and testing on the Cobas 6800, 5/10 (50.0%) RT-PCR negative samples, with paired positive ID NOW swabs, resolved as positive. One sample was unable to undergo additional testing.

PPA between the ID NOW and RT-PCR, stratified based on individual characteristics, is provided in Figure 1. The highest ID NOW PPA (98.2%) was in individuals with symptoms ≤ 7 days and who had the ID NOW performed within an hour, followed by individuals with symptoms ≤ 7 days (91.2%), and in samples with ID NOW performed within an hour (89.8%). Further details are provided within the supplementary material.

**Figure 1:**
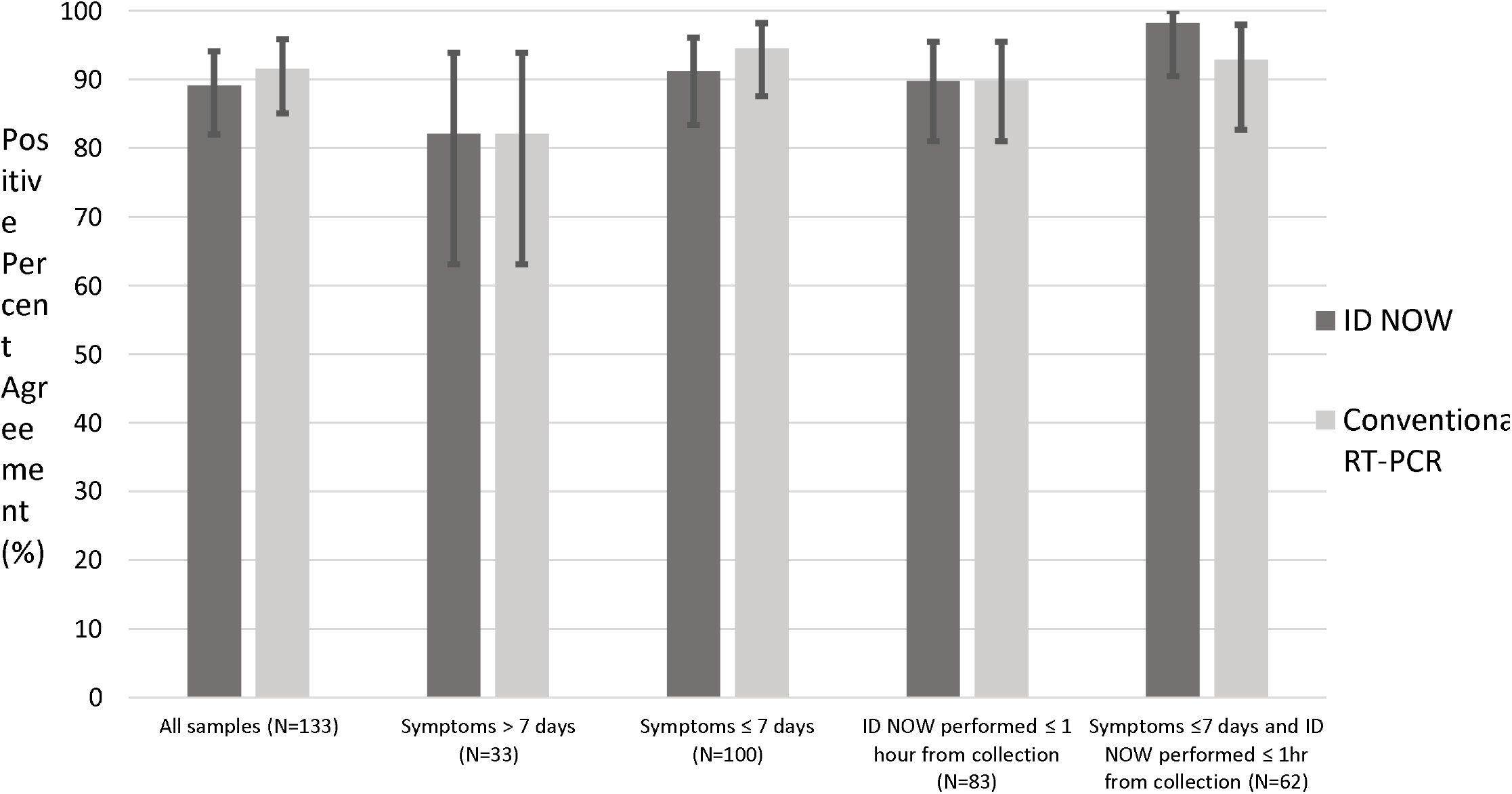
Positive percent agreement of the ID NOW and RT-PCR from samples, stratified based on sample/individual characteristics, with 95% confidence intervals provided. Stratified samples include those among all individuals (N=133), individuals with symptoms > 7 days (N=33), individuals with symptoms ≤ 7 days (N=100), individuals who had the ID NOW performed within ≤1 hour from collection (N=83), and individuals with symptoms ≤ 7 days and ID NOW performed ≤1 hour from collection (N=62).

Twenty asymptomatic individuals at low risk of COVID-19 were tested, all of which were negative on the ID NOW and RT-PCR. All four retrospective samples positive for SARS-CoV-2 were positive on the ID NOW. All 11 retrospective samples containing other respiratory viruses tested were negative. These samples were previously positive for one of either human metapneumovirus, adenovirus, parainfluenza virus, other coronavirus (NL63, HKU1, NL63), enterovirus, respiratory syncytial virus, influenza A H3N2, influenza A H1N1, or influenza B.

## DISCUSSION

Our study compared the PPA of the ID NOW to RT-PCR among individuals confirmed to have SARS-CoV-2 infection. Overall, PPA of the ID NOW instrument was high at 89.9% and comparable to our laboratory’s RT-PCR (91.6%) for this population. In the cohort of patients with symptom onset ≤ 7 days prior to collection, and whose samples could be tested within one hour from collection, the PPA increased to 98.2%. While there were instances in our study where the ID NOW was positive and the RT-PCR was negative, we believe these are true positives based on several reasons. Participants recruited in our study were all recently diagnosed with COVID-19; none of the samples from the asymptomatic individuals at low risk of COVID-19 gave false positive results throughout the study; when retested in triplicate or on an alternative platform, 40% of these samples had detectable RNA present; and no issues with false positive results have been identified by the ID NOW manufacturer or among previous publications within the literature.^2-17^

PPA of the ID NOW varies widely in the literature from 48.0% to 94.1%.^2-17^ However, most of these studies suffered from poor study design or were conducted when the manufacturer still considered placing swabs in UTM as appropriate for sample collection (prior to April, 2020). Several studies did not adhere to the recommended time limit of 1 hour for time of collection to result which, as confirmed in this study, is a statistically significant factor in ID NOW’s performance.^2,11,15^ One of the major studies to discredit the performance of the ID NOW compared dry nasal swabs (tested on the ID NOW) to nasopharyngeal swabs (tested on Cepheid Xpert Xpress), which is an inappropriate comparison given the superiority of positivity rate among nasopharyngeal specimens to nasal specimens.^19,21^ Furthermore, Basu *et al*. tested patients with symptom onset up to 1 month from time of sample collection, and it is unclear how many patients with confirmed COVID-19 had symptoms ≤ 7 days.^3^ Among the studies that adhered to current ID NOW manufacturer recommendations for testing, ID NOW PPA was 66.7% and 94.1%.^16,17^ However, both studies had small sample sizes (<20 samples positive for SARS-CoV-2).

Limitations of our study include the low number of hospitalized patients recruited such that we cannot make strong conclusions about the ID NOW performance among this population. The majority of throat swabs tested on the ID NOW were collected before the comparator swab (78.0%). However, we did not observe any difference in ID NOW or RT-PCR positivity rate when comparing patients who had ID NOW throat swab collected first vs second (Table 5 and supplementary material). There were discrepancies between ID NOW positive, RT-PCR negative specimens, that could have resulted from multiple factors, such as intra-collector variability in the throat swab collections and degradation of virus during transportation/storage, as opposed to false positive ID NOW results.

The strengths of our study include the large number of COVID-19 positive individuals recruited, particularly those residing within the community. We adhered to the manufacturer’s recommendations, as much as possible, to recruit COVID-19 positive individuals with symptoms ≤ 7 days at time of collection and to test samples on the ID NOW within an hour from collection. Adhering to these requirements was difficult, as it often took several days from symptom onset for an individual in the community to get swabbed and test results reported. Consequently, the symptom onset was often near seven days. Meeting the one-hour criteria was challenging as we had to drive back and forth from participants’ households to our laboratory, and many participants were located in distant parts of the city (e.g. >30 min drive to the laboratory) and other obstacles (e.g. traffic) increased transit time. Another strength of our study was the concurrent testing of asymptomatic individuals at low risk of COVID-19, and retrospective samples positive for other respiratory viruses, throughout the study to ensure there were no issues with false positive results (e.g. caused by contamination or cross-reactivity).

## CONCLUSIONS

In conclusion, the ID NOW was found to be a comparable method to our RT-PCR for the detection of SARS-CoV-2 among individuals with symptomatic COVID-19 infection. PPA was enhanced when tested on individuals with symptom onset ≤ 7 days and when time from collection to testing was within one hour. These results reassure us that the ID NOW is a reliable test method in symptomatic individuals, especially when adhering to the FDA-approved indications and recommendations for testing. Given the speed and low-complexity of ID NOW testing, these instruments can truly be used as a point of care device. As such, they will play an impactful role in combating the COVID-19 pandemic by improving testing in settings where low-volume rapid (<1h) turnaround times are much needed, such as among difficult to reach populations (e.g. homeless), inpatients with suspected nosocomial infection, and in rural areas where access to a laboratory is limited because transportation delays are significant.

## Data Availability

Available upon request

## Acknowledgments

This work was funded using internal operating funds of Alberta Precision Laboratories and Alberta Health Services. Test kits and instruments were paid for by the Public Health Agency of Canada. We thank the AHS mobile integrated health team for collecting samples in the community and Alberta Precision Laboratory staff for assistance with testing of samples.

