## Supplementary Material for "Acceptable Performance of the Abbott ID NOW Among Symptomatic Individuals with Confirmed COVID-19"

Table 1: Details on the ID NOW positive, RT-PCR negative results (N=10)

| **Number** | **Result when tested in triplicate (lab developed test)** | **Result when tested in triplicate (CDC method)** | **Results when tested on Cobas 6800** | **Duration of symptoms at time of collection** | **Symptoms** |
| --- | --- | --- | --- | --- | --- |
| 1 | U/U/U | U/U/U | U/U | 8* | Shortness of breath |
| 2 | U/U/U | U/U/U | U/U | 12* | Shortness of breath |
| 3 | U/U/U | U/U/U | U/U | 4 | Nasal congestion, anosmia, ageusia |
| 4 | U/40.0/U | U/36.5/37.2 | 33.6/35.7 | 14 | Nausea, malaise |
| 5 | U/U/U | 35.78/36.87/35.82 | N/A | 4 | Nasal congestion, cough, anosmia, ageusia |
| 6 | U/U/U | U/U/U | U/U | 11 | Fever, cough, anosmia, ageusia, pharyngitis |
| 7 | U/37.9/U | 37.8/38.1/U | N/A | 7 | Rhinorrhea, cough, anosmia |
| 8 | U/36.8/U | 36.7/U/U | 32.3/34.4 | 5 | Headache, myalgia, malaise |
| 9 | U** | N/A | N/A | 4 | Pharyngitis, headache |
| 10 | U/37.9/38.1 | U/U/U | N/A | 17 | Chest pain |

U = undetectable

Numbers- Ct values where applicable

*Hospitalized

**Additional testing unable to be performed due to specimen being accidentally discarded

Table 2: Characteristics between RT-PCR negative and RT-PCR positive samples.

|  | **RT-PCR negative (N=24)** | **RT-PCR positive (N=106)** | **P-value** |
| --- | --- | --- | --- |
| **Mean symptoms (days)** | 7.8 | 6.8 | 0.034 |
| **Symptoms** ≤ **7 days** | 58.3% | 79.2% | 0.032 |
| **Mean age (years)** | 46.7 | 42.3 | 0.236 |
| **Throat swab tested on ID NOW collected first** | 79.2 | 78.1 | 0.909 |
| **Hospitalized** | 16.7% | 7.5% | 0.163 |
| **Male gender** | 45.8% | 34.9% | 0.316 |

Table 3: Comparison of results from the ID NOW and RT-PCR in COVID-19 patients without

symptoms (N=14)

|  |  | **RT-PCR** | |
| --- | --- | --- | --- |
|  |  | Positive | Negative |
| **ID NOW** | Positive | 7 | 1 |
|  | Negative | 1 | 5 |

Table 4: Positive percent agreement (PPA) between ID NOW and RT-PCR in asymptomatic

COVID-19 patients (N=14). PPA calculated assuming any positive is a true positive.

|  | **Positive percent agreement** |
| --- | --- |
| **ID NOW** | 88.9% [51.8% - 99.7%] |
| **Conventional RT-PCR** | 88.9% [51.8% - 99.7%] |

Table 5: Comparison of the ID NOW and RT-PCR in hospitalized COVID-19 patients (N=12)

|  |  | **RT-PCR** | |
| --- | --- | --- | --- |
|  |  | Positive | Negative |
| **ID NOW** | Positive | 7 | 2 |
|  | Negative | 1 | 2 |

Table 6: Positive percent agreement (PPA) between ID NOW and RT-PCR in

Hospitalized COVID-19 patients (N=14). PPA calculated assuming

any positive is a true positive.

|  | **Positive percent agreement** |
| --- | --- |
| **ID NOW** | 90.0% [55.5% - 99.8%] |
| **Conventional RT-PCR** | 80.0% [44.4% - 97.5%] |

Table 7: Comparison of the ID NOW and RT-PCR in COVID-19 patients with symptoms > 7 days

(N=33)

|  |  | **RT-PCR** | |
| --- | --- | --- | --- |
|  |  | Positive | Negative |
| **ID NOW** | Positive | 18 | 5 |
|  | Negative | 5 | 5 |

Table 8: Positive percent agreement (PPA) between ID NOW and RT-PCR in COVID-19 patients

with symptoms > 7 days (N=33). PPA calculated assuming any positive is a true positive.

|  | **Positive Percent Agreement** |
| --- | --- |
| **ID NOW** | 82.1% [63.1% - 93.9%] |
| **conventional RT-PCR** | 82.1% [63.1% - 93.9%] |

Table 9: Comparison of the ID NOW and RT-PCR in COVID-19 patients with symptoms ≤ 7 days

(N=100)

|  |  | **RT-PCR** | |
| --- | --- | --- | --- |
|  |  | Positive | Negative |
| **ID NOW** | Positive | 78 | 5 |
|  | Negative | 8 | 9 |

Table 10: Positive percent agreement (PPA) -19 patients with symptoms ≤ 7 days (N=100). PPA

calculated assuming any positive is a true positive.

|  | **Positive Percent Agreement** |
| --- | --- |
| **ID NOW** | 91.2% [83.4% - 96.1] |
| **RT-PCR** | 94.5% [87.6% - 98.2%] |

Table 11: Comparison of the ID NOW and RT-PCR in COVID-19 Individuals who had the ID NOW

tested within 1 hour (N=83)

|  |  | **RT-PCR** | |
| --- | --- | --- | --- |
|  |  | Positive | Negative |
| **ID NOW** | Positive | 63 | 8 |
|  | Negative | 8 | 4 |

Table 12: Positive percent agreement (PPA) between ID NOW and RT-PCR in

COVID-19 Individuals who had the ID NOW tested within 1 hour (N=83). PPA calculated

assuming any positive is a true positive.

|  | **Positive percent agreement** |
| --- | --- |
| **ID NOW** | 89.9% [81.0% - 95.5%] |
| **RT-PCR** | 89.9% [81.0% - 95.5%] |

Table 13: Two by two table between ID NOW and RT-PCR in COVID-19 Individuals with

symptoms ≤ 7 days and ID NOW test conducted within an hour from collection (N=62)

|  |  | **RT-PCR** | |
| --- | --- | --- | --- |
|  |  | Positive | Negative |
| **ID NOW** | Positive | 50 | 4 |
|  | Negative | 1 | 6 |

Table 14: Positive percent agreement (PPA) between ID NOW and RT-PCR in COVID-19

Individuals with symptoms ≤ 7 days and ID NOW test conducted within an hour from

collection (N=62). PPA calculated assuming any positive is a true positive.

|  | **Positive percent agreement** |
| --- | --- |
| **ID NOW** | 98.2% [90.5% – 100%] |
| **RT-PCR** | 92.9% [82.7% - 98.0] |
